# Regional replacement of SARS-CoV-2 variant BA.1 with BA.2 as observed through wastewater surveillance

**DOI:** 10.1101/2022.04.22.22274160

**Authors:** Alexandria B. Boehm, Bridgette Hughes, Marlene K. Wolfe, Bradley J. White, Dorothea Duong, Vikram Chan-Herur

**Affiliations:** Department of Civil & Environmental Engineering, Stanford University, 473 Via Ortega, Stanford, CA, USA, 94305; Verily Life Sciences, South San Francisco, CA, USA, 94080; Gangarosa Department of Environmental Health, Rollins School of Public Health, Emory University, 1518 Clifton Rd, Atlanta, GA, USA, 30322

**Keywords:** Omicron, wastewater, SARS-CoV-2, wastewater-based epidemiology

## Abstract

An understanding of circulating SARS-CoV-2 variants can inform pandemic response, vaccine development, disease epidemiology, and use of monoclonal antibody treatments. We developed custom assays targeting characteristic mutations in SARS-CoV-2 variants Omicron BA.1 and BA.2 and confirmed their sensitivity and specificity in silico and *in vitro*. We then applied these assays to daily wastewater solids samples from eight publicly owned treatment works in the greater Bay Area of California, USA, over four months to obtain a spatially and temporally intensive data set. We documented regional replacement of BA.1 with BA.2 in agreement with, and ahead of, clinical sequencing data. This study highlights the utility of wastewater surveillance for real time tracking of SARS-CoV-2 variant circulation.

**Synopsis:** Wastewater surveillance was used to document regional emergence of SARS-CoV-2 variant Omicron BA.2 ahead of clinical surveillance.

**Graphical Abstract:** 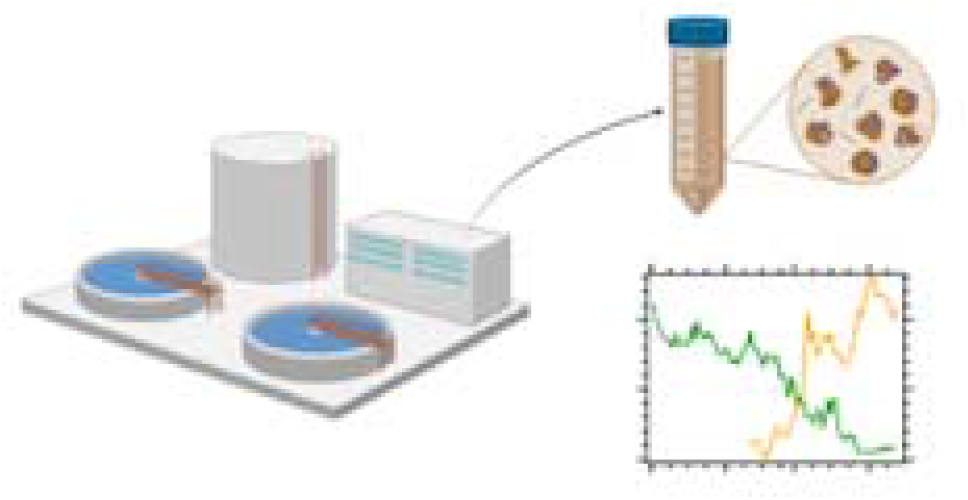

## Introduction

Knowing when a new SARS-CoV-2 variant arrives locally is of utmost importance to pandemic response in order to ensure a rapid response to new features of the virus. In late 2021, the Omicron variant emerged in South Africa and quickly spread to Europe and then North America during a period of time during which Delta was the dominant circulating variant. The Omicron variant resulted in a virus more transmissible and less susceptible to therapeutics and vaccines than previous lineages^1^. Given these implications for public health, timely identification of its introduction into a new community was critical. Variant identification is usually accomplished through sequencing of clinical specimens, but the time between specimen collection, and the return of sequencing information to public health officials is approximately two weeks. For example, outbreak.info^2^, a website that provides analysis and visualization of SARS-CoV-2 clinical sequences deposited in GISAID^3,4^, stresses data are unreliable and incomplete during the 1-2 week window preceding the query date.

Wastewater represents a biological composite sample from the contributing community. It contains feces, urine, sputum, mucus, blood, vomitus, and any other excretion that goes down a drain into the sewer network. Global investigations have illustrated that SARS-CoV-2 RNA concentrations, whether measured by RT-QPCR or digital RT-PCR, or in the liquid or solid phase of wastewater, correlates well with laboratory-confirmed incident COVID-19 cases in the associated sewersheds^5–8^. A number of studies have illustrated that circulating or novel variants can be identified in wastewater using RT-PCR assays that target a characteristic variant mutation^9–11^ and by sequencing SARS-CoV-2^12^. We previously developed targeted RT-PCR assays for characteristic mutations in Alpha, Mu, Lambda, Delta, and Omicron BA.1, and showed that the concentrations of these mutations, normalized by a pan-SARS-CoV-2 N gene target is strongly correlated to the fraction of clinical specimens classified as the associated variant^10,13^ at two publicly owned treatment works (POTWs). Here, we extend that previous work by developing and testing two new assays, one that targets characteristic mutations in the BA.2 sublineage of Omicron and another that targets a set of mutations present in both BA.1 and BA.2, and then applying them to wastewater samples collected daily at eight POTW in the greater Bay Area, California, USA. Results presented here were available publicly within 24 hours of sample collection via our website (wbe.stanford.edu), and these data allowed us to observe near real-time regional replacement of BA.1 with BA.2.

## Materials and Methods

### Characteristic spike mutation assay development

Digital RT-PCR assays were designed to target the BA.2 characteristic mutation LPPA24S (a 9 bp deletion), and a region of 5 adjacent SNPs common to BA.1 and BA.2 (S477N,T478K,Q493R,Q498R,Y505H, hereafter S:477-505). The del143-145 assay targeting a mutation present in BA.1 was developed in a similar manner described elsewhere^10^. According to sequences deposited in GISAID and accessed via outbreak.info (accessed 16 April 2022), the LPPA24S mutation is present in 92% of BA.2 genomes, including sublineages, (hereafter BA.2*) and 0.003% of BA.1 genomes, including sublineages, (hereafter BA.1*) globally whereas the del143-145 mutation is present in 93% BA.1* and 0% BA.2* genomes, globally (Table S1). Assays were developed *in silico* using Primer3Plus (https://primer3plus.com/) (Table S2).

Primers and probe sequences (Table 1) were screened for specificity *in silico* using NCBI BLAST, and then tested *in vitro* against various respiratory viruses and gRNA from wild-type (WT) SARS-CoV-2 and its variants (see SI). Viral RNA was used undiluted as template in digital droplet RT-PCR with mutation primer and probes. The concentration of gRNA used for in vitro specificity testing was approximately 275 copies per well.

**Table 1.**
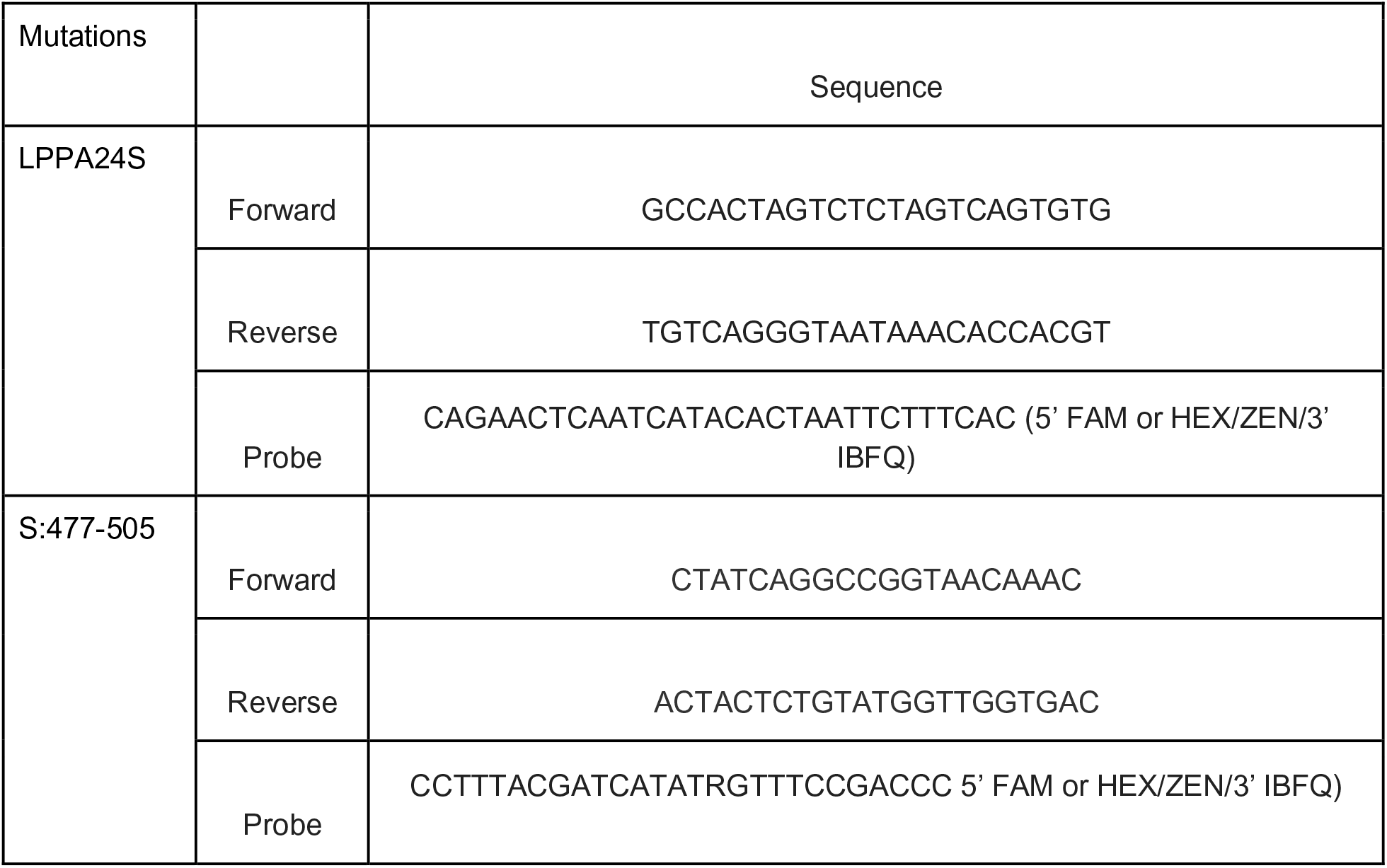
Primers and probes for new mutation assays. Primers and probes for other assays that are previously published are in Table S5. Information on the fluorescent molecule and quenchers used for the probes are provided in parentheses after their sequence. FAM, 6-fluorescein amidite; HEX, hexachloro-fluorescein; ZEN, a proprietary internal quencher from IDT; IBFQ, Iowa Black FQ.

The sensitivity and specificity of the mutation assays were further tested by diluting target variant gRNA (Table S1) in no (0 copies), low (100 copies), and high (10,000 copies) background of WT SARS-CoV-2 gRNA.

### Wastewater samples

Eight POTWs serving populations of the Greater Bay Area and Sacramento, California, USA were included in the study. They serve between 66,000 and 1.5 million people; further descriptions are elsewhere.^14^

50□mL of settled solids was collected each day from each POTW between 1 January 2022 and 12 or 13 April 2022, during the period of time when Omicron BA.2 emerged in the region. Detailed sample collection methods are provided elsewhere^14^ and in the SI. Samples were immediately stored at 4°C and transported to the laboratory by a courier service where processing began within 6□h of collection. Between 94 and 103 samples from each POTW were included in the analysis (total of 810).

RNA was extracted from 10 replicate aliquots of each dewatered settled solids sample and PCR inhibitors were removed^14^ (see SI). RNA was processed immediately to measure concentrations of the N gene of SARS-CoV-2, the BA.1* characteristic mutation del143-145^10^, the BA.2* characteristic mutation (LPPA24S), pepper mild mottle virus (PMMoV), and bovine coronavirus (BCoV) recovery using digital droplet RT-PCR methods^14,15^. The N gene target is present in all variants and is a pan-SARS-CoV-2 gene target. PMMoV is highly abundant in human stool and wastewater globally^16,17^ and is used as an internal recovery for the wastewater samples^18^. BCoV was spiked into the samples and used as an additional recovery control; all samples were required to have greater than 10% BCoV recovery. RNA extraction and PCR negative and positive controls were included to ensure no contamination. Extracted RNA samples from one POTW (SJ) were then stored at −80°C between 1 and 7 days before they were analyzed for S:477-505 and the N gene. Each of the 10 replicate RNA extracts was run in its own well, and the 10 wells were merged for analysis. Wastewater data are available publicly at the Stanford Digital Repository (https://doi.org/10.25740/cf848zx9249); results below are reported as suggested in the EMMI guidelines^19^.

### ddRT-PCR

The ddRT-PCR methods applied to wastewater solids to measure PMMoV and BCoV are provided in detail elsewhere^14^. The N gene and del143-145 mutation were multiplexed along with an S gene assay (not reported herein). LPPA24S mutation was multiplexed in an assay along with an influenza A and RSV N gene target (only LPPA24S reported herein). The S:477-505 mutation was multiplexed with the N gene and a characteristic mutation in Delta (not reported). Each 96-well PCR plate of wastewater samples included PCR and extraction positive and negative controls. Additional details of the PCR methods are provided in the SI.

QuantaSoft™ Analysis Pro Software (Bio-Rad, version 1.0.596) was used for thresholding. In order for a sample to be recorded as positive, it had to have at least three positive droplets. For the wastewater solid samples, three positive droplets across ten merged wells corresponds to a concentration between ∼500-1000 copies per g (cp/g); the range in values is a result of the range in the equivalent mass of dry solids added to the wells.

Concentrations of RNA targets were converted to concentrations per dry weight of solids in units of copies/g dry weight using dimensional analysis. Dry weight of dewatered solids was determined by oven drying^20^. The total error is reported as standard deviations and includes the errors associated with the Poisson distribution and the variability among the 10 replicates.

### Variants present in regional clinical specimens

The 7-d, centered, rolling average proportion of clinical specimens sequenced from California classified as Omicon BA.1* and BA.2* as a function of specimen collection data were acquired from GISAID^3,4^ on 21 April 2022.

### Statistics

We normalized mutation concentrations by N gene concentrations to represent the fraction of total SARS-CoV-2 RNA (represented by the N gene assay target which is conserved across variants) that comes from the variant; hereafter referred to as relative mutation concentration. We used Kendall’s tau to test for associations between the 5-d trimmed smoothed average relative concentrations and the proportion of clinical specimens assigned to the corresponding variants as the two variables were not normally distributed (Shaprio Wilk test, p<0.05 for all). The daily measured relative concentration was matched to the 7-d, centered, rolling average fraction of clinical specimens classified as the associated variant obtained from GISAID. All analysis was carried in R Studio version 1.4.1106.

## Results and Discussion

### Variant mutation assay specificity

*In silico* analysis indicated no cross reactivity between LPPA24S (targeting BA.2*) and S:477-505 (targeting BA.1* and BA.2*) assays and deposited sequences in NCBI. When challenged against a respiratory virus panel and various non-target SARS-CoV-2 variant gRNA no cross reactivity was observed; the assays amplified gRNA from BA.2, and BA.1 and BA.2, respectively. Positive controls and NTCs run on sample plates were positive and negative. When target gRNA was diluted into a background of low and high non-target gRNA from WT SARS-CoV-2; measurements were similar when there was no, low, or high background gRNA (Figure S1). Results indicate the new mutation assays are specific and sensitive. Wolfe et al.^10^ showed the del143-145 (targeting BA.1) assay was sensitive and specific.

### Variant RNA concentrations in wastewater solids

All positive and negative controls were positive and negative, respectively, indicating assays performed well and without contamination. BCoV recoveries were higher than 10% and PMMoV concentrations within the expected range for the POTW suggesting an efficient and acceptable recovery of RNA during RNA extraction (Figure S2).

Across all eight POTWs a common pattern in N, del143-145, and LPPA24S concentrations is observed. Between 1 Jan and late Feb to mid-March 2022, N and del143-145 concentrations are approximately the same, and both decrease from 10^6^ cp/g to 10^4^ cp/g (Figure 1). The two order of magnitude decrease is coincident with a decline in laboratory-confirmed incident COVID-19 cases in the sewersheds after the winter Omicron surge in the region (case data not shown). In late Feb to mid-March until mid-April, both N and LPPA24S concentrations increase from 10^3^ cp/g to 10^5^ cp/g, while del143-145 concentrations decrease from 10^4^ cp/g to 10^2^ cp/g or non-detect. LPPA24S concentrations are similar to those of N at the POTWs during that same time period.

**Figure 1.**
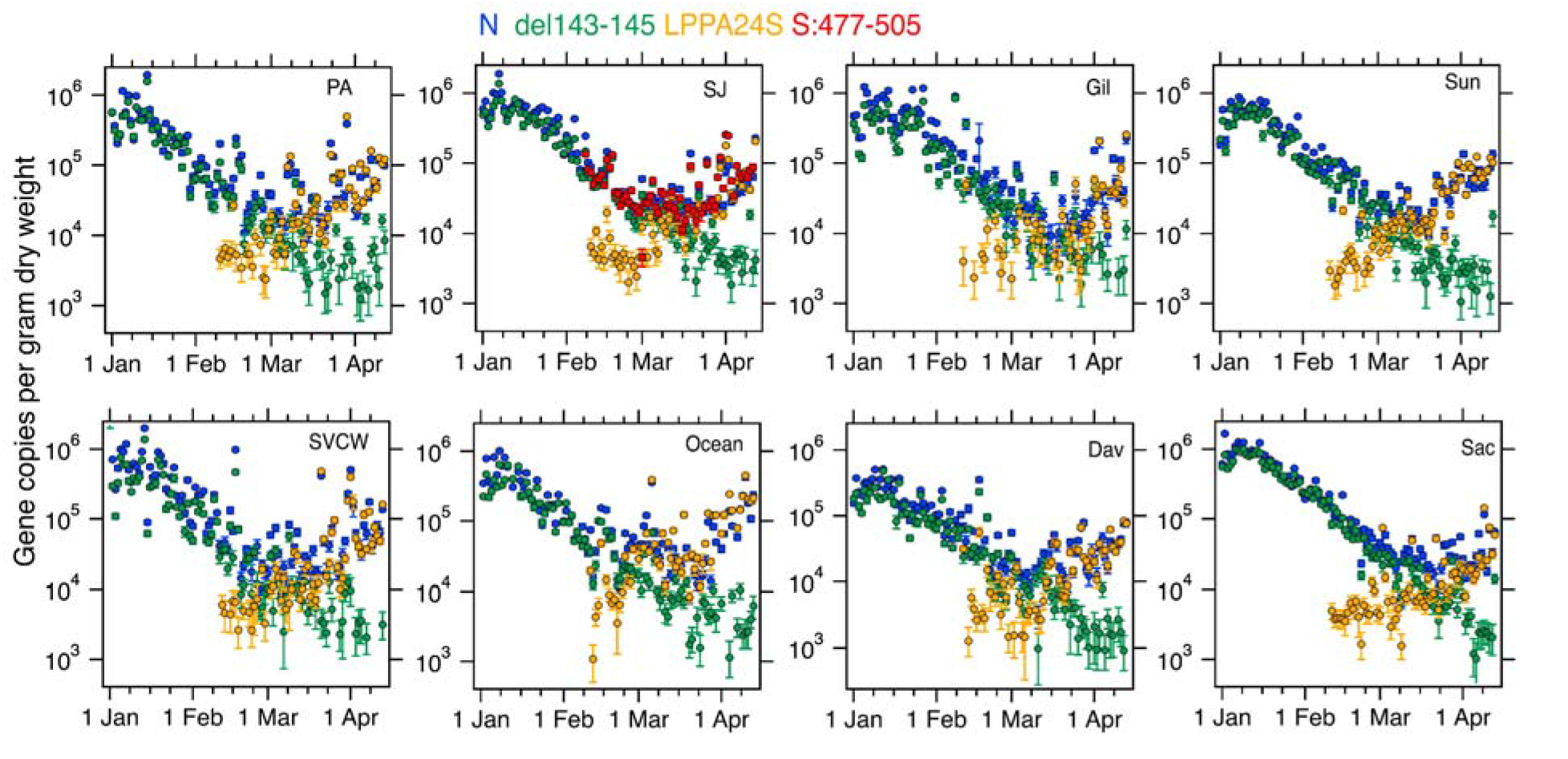
Concentrations of N, del143-145, LPPA24S, and S:477-505 at 8 POTWs in the study (name indicated in corner). Error bars are 68% confidence intervals that include error from variation among 10 replicate wells, and Poisson error (“total error” as reported by the instrument). If error bars cannot be seen, they are smaller than the symbol.

The ratios del143-145/N and LPPA24S/N describe the concentrations of the mutations in relation to a conserved gene target. As a mutation’s relative concentration approaches 1, it suggests that the variant associated with the mutation may be dominant in the wastewater. Del143-145 relative concentration was a maximum and close to 1 at most POTWs at the beginning of the time series (1 January 2022) and then fell to nearly 0 by mid-April 2022 (Figure 2). At the same time, LPPA24S relative concentration increased from 0 when it was first measured to near 1 at all POTWs. Depending on POTW, the date on which the LPPA24S relative concentration surpassed that of del143-145 ranged from 26 February to 19 March (Table S3). This date was earliest (26 Feb) at Ocean, located in San Francisco, and latest at Gil and Sac; the date is positively correlated with the distance from Ocean (r =0.9, p<0.05, n=8) suggesting a spatial pattern in BA.2* emergence. Regional replacement of BA.1* with BA.2* appears to be complete at all POTWs by the end of the times series as the del143-145 relative concentrations are close to 0 and LPPA24S relative concentrations approach 1. At times, relative concentrations may exceed 1; this may be due to variability in the performance of the digital RT-PCR assays. It is important to note that the relative concentrations do not incorporate measurement error of the numerator or denominator.

**Figure 2.**
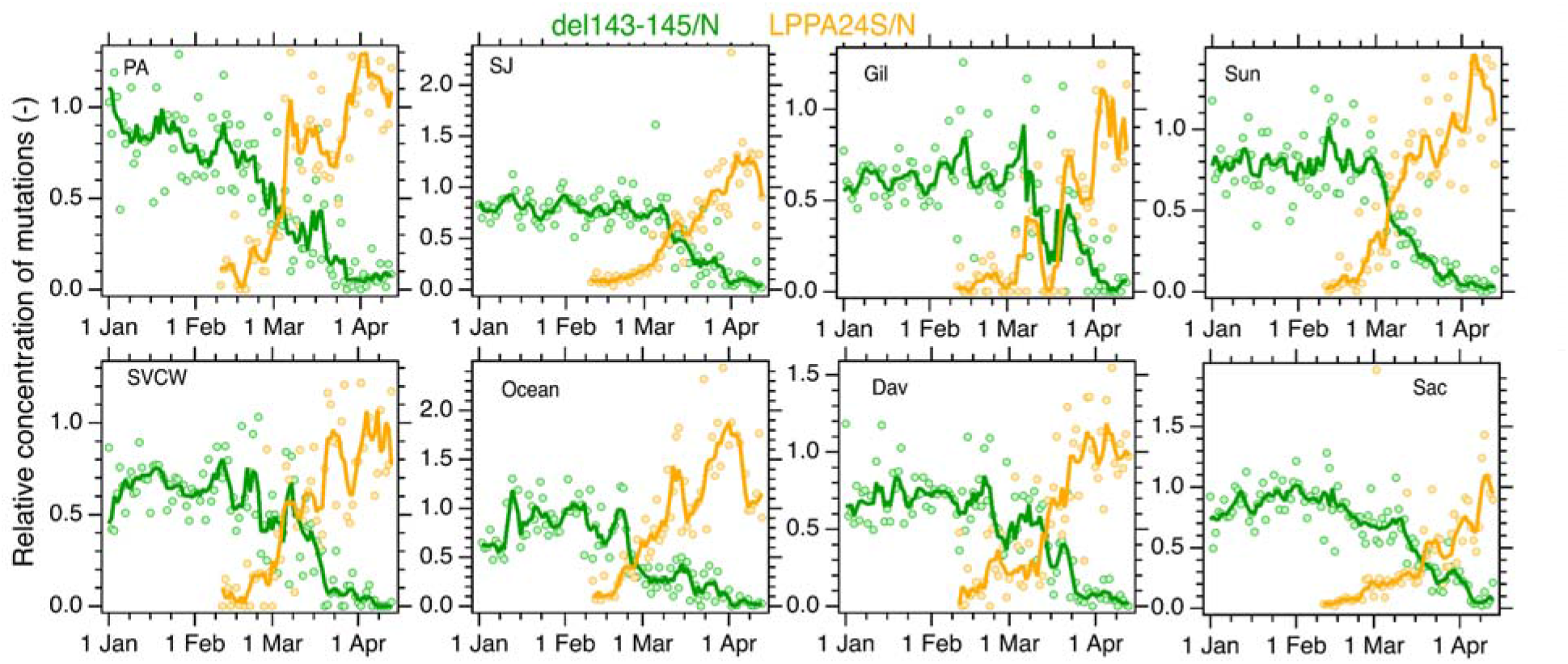
Relative concentrations of del143-145 and LPPA24S (their concentrations normalized by the N gene) at 8 POTWs in the study (name indicated in corner). Markers are raw ratio values. Lines represent 5-day trimmed moving averages.

At SJ, we measured the concentration of an additional set of adjacent mutations present in both BA.1 and BA.2 (S:477-505) (Figure 1). S:477-505 concentrations agree well with the N concentrations during a period of time when BA.1* was dominant (prior to mid-March) and when BA.2* was dominant (thereafter).

del143-145 and LPPA24S relative concentrations at each POTW are positively and significantly correlated with the fraction of clinical cases in California assigned as BA.1* and BA.2*, respectively (Table S4). Tau between relative del143-145 concentrations and proportion of cases assigned as BA.1* ranged from 0.51 at Gil to 0.81 at SAC (median tau = 0.68, all p<10^−7^, n = 94-103). Tau between relative LPPA24S concentrations and proportion of cases assigned as BA.2* ranged from 0.67 at Gil to 0.90 at SAC (median tau = 0.75, all p<10^−5^, n = 58-62). Tau values were similar when calculated using raw relative concentrations (see SI). State level clinical data suggest proportion of cases caused by BA.2* exceeds that of BA.1* after 21 March (Figure 3), later than any wastewater suggests this occurred regionally. Ideally, these analyses should be carried out using data on clinical variants in the sewersheds of the eight POTWs, but such data is not readily available, so we had to rely on state-level data.

**Figure 3.**
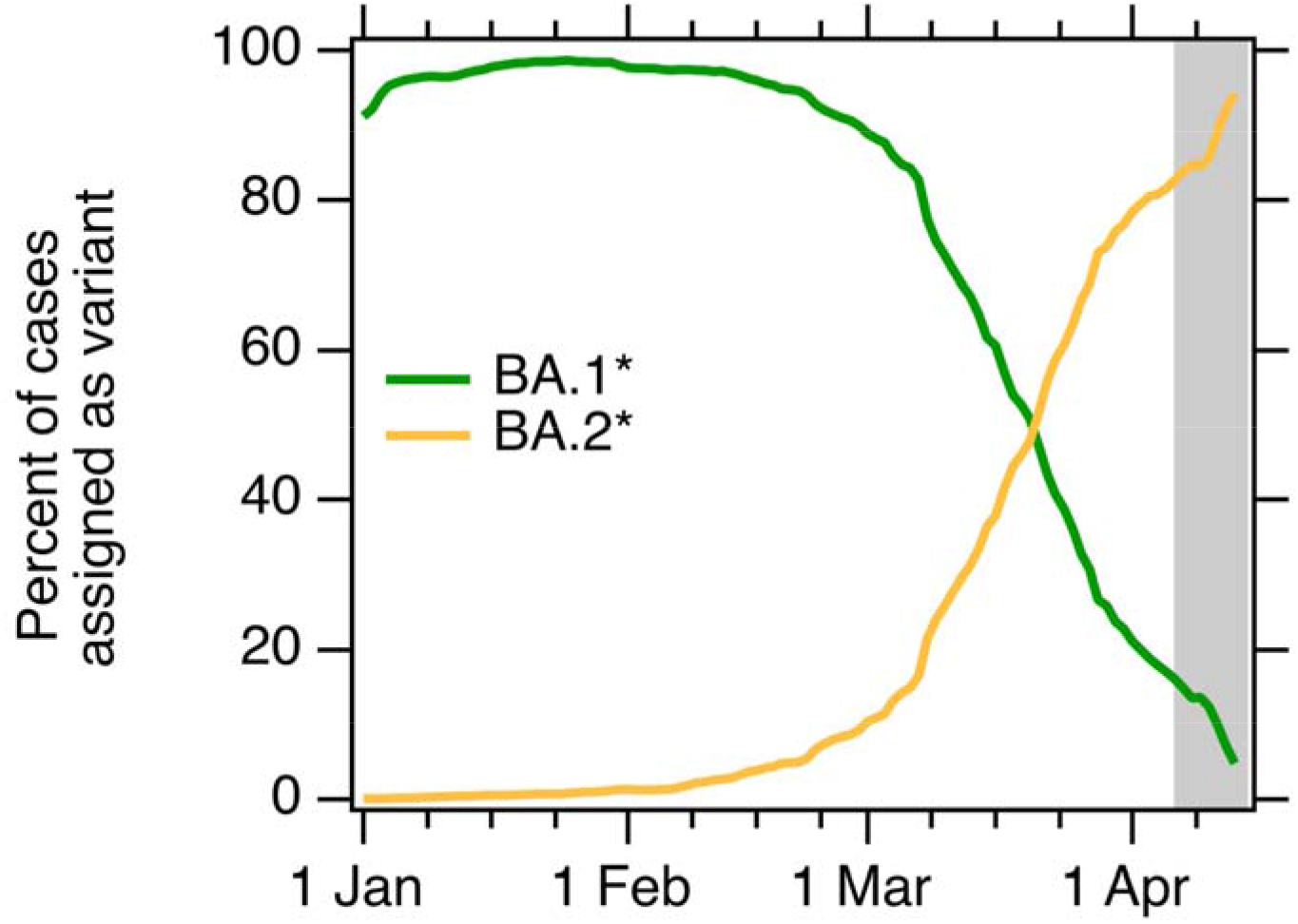
Percentage of clinical cases in California classified as indicated BA.1* and BA.2* as a function of specimen collection date; data represent 7-d moving average as acquired from GISAID. Data available through 13 April when downloaded from GISAID on 21 April 2022. Gray area represents the time period for which data were incomplete on 14 April 2022, when wastewater measurements for the last date in our data series (12 or 13 April 2022) were available.

State-level clinical sample sequencing data on variant occurrence is typically 1-2 weeks delayed owing to the time required for laboratory processing and reporting. In addition, there can be large uncertainties in the proportion estimates preceding the data for which the most recent data are available. This is illustrated in Figure 3 where we highlight the approximate time period for which data were incomplete on 14 April 2022, when wastewater measurements for the last date in our data series (12 or 13 April 2022) were available publicly on our website (wbe.stanford.edu). During this same time period, wastewater data suggest BA.2* is near 100% and BA.1* is near 0%, which a week later (when data were downloaded from GISAID on 21 April 2022) was confirmed by clinical sequencing data. This highlights an important advantage of using wastewater surveillance for variant tracking.

Real-time estimates of variant abundance with high spatial resolution, such as those from wastewater surveillance, could allow clinicians and regulators to identify the most effective treatment for a particular community. The monoclonal antibody therapy sotrovimab provides a recent and practical example. Sotrovimab was granted emergency use authorization by the US FDA for treatment of infection by variants including BA.1, but not BA.2. As BA.2 took over, its use was discontinued by multi-state HHS region blocks, informed by clinical sequencing data. While sotrovimab was allowed for use in California until March 30, ourdata suggest BA.2 became the dominant variant in the Bay Area weeks prior, but before BA.2 had taken over all of California and HHS region 9. Real-time wastewater variant estimates could have allowed the drug’s use to be targeted spatially and temporally to maximize effectiveness.

## Data Availability

All data are available through Stanford Digital Repository: https://doi.org/10.25740/cf848zx9249

## Acknowledgements

This work is supported by the CDC Foundation.Numerous people contributed to sample collection and case data acquisition, including Srividhya Ramamoorthy (Sac), Michael Cook (Sac), Ursula Bigler (Sac), James Noss (Sac), Lisa C. Thompson (Sac), Payak Sarkar (SJ), Noel Enoki (SJ), and Amy Wong (SJ), Lily Chan (Ocean), the Oceanside plant operations personnel, Karin North (PA), Armando Guizar (PA), Saeid Vaziry (Gil), Chris Vasquez (Gil), Alo Kauravlla (Sun), Maria Gawat (SVCW), Tiffany Ishaya (SVCW), and Jeromy Miller (Dav). This study was performed on the ancestral and unceded lands of the Muwekma Ohlone people. We pay our respects to them and their Elders, past and present, and are grateful for the opportunity to live and work here. The graphic abstract was made using Biorender (biorender.com).

## Supporting Information

Additional methods, Tables S1-S5, Figures S1-S3. This paper has been previously submitted to medRxiv, a preprint server for Health Sciences. The preprint can be cited as: XXXX (to be provided).

## Competing Interests

BH, DD, VC-H and BW are employees of Verily Life Sciences.

## Notes

### Funding Statement

This study was funded by CDC-Foundation.

### Summary of Updates

Corrected author list.

## References

(1) Viana, R.; Moyo, S.; Amoako, D. G.; Tegally, H.; Scheepers, C.; Althaus, C. L.; Anyaneji, U. J.; Bester, P. A.; Boni, M. F.; Chand, M.; Choga, W. T.; Colquhoun, R.; Davids, M.; Deforche, K.; Doolabh, D.; du Plessis, L.; Engelbrecht, S.; Everatt, J.; Giandhari, J.; Giovanetti, M.; Hardie, D.; Hill, V.; Hsiao, N.-Y.; Iranzadeh, A.; Ismail, A.; Joseph, C.; Joseph, R.; Koopile, L.; Kosakovsky Pond, S. L.; Kraemer, M. U. G.; Kuate-Lere, L.; Laguda-Akingba, O.; Lesetedi-Mafoko, O.; Lessells, R. J.; Lockman, S.; Lucaci, A. G.; Maharaj, A.; Mahlangu, B.; Maponga, T.; Mahlakwane, K.; Makatini, Z.; Marais, G.; Maruapula, D.; Masupu, K.; Matshaba, M.; Mayaphi, S.; Mbhele, N.; Mbulawa, M. B.; Mendes, A.; Mlisana, K.; Mnguni, A.; Mohale, T.; Moir, M.; Moruisi, K.; Mosepele, M.; Motsatsi, G.; Motswaledi, M. S.; Mphoyakgosi, T.; Msomi, N.; Mwangi, P. N.; Naidoo, Y.; Ntuli, N.; Nyaga, M.; Olubayo, L.; Pillay, S.; Radibe, B.; Ramphal, Y.; Ramphal, U.; San, J. E.; Scott, L.; Shapiro, R.; Singh, L.; Smith-Lawrence, P.; Stevens, W.; Strydom, A.; Subramoney, K.; Tebeila, N.; Tshiabuila, D.; Tsui, J.; van Wyk, S.; Weaver, S.; Wibmer, C. K.; Wilkinson, E.; Wolter, N.; Zarebski, A. E.; Zuze, B.; Goedhals, D.; Preiser, W.; Treurnicht, F.; Venter, M.; Williamson, C.; Pybus, O. G.; Bhiman, J.; Glass, A.; Martin, D. P.; Rambaut, A.; Gaseitsiwe, S.; von Gottberg, A.; de Oliveira, T. Rapid Epidemic Expansion of the SARS-CoV-2 Omicron Variant in Southern Africa. Nature 2022, 603 (7902), 679–686. https://doi.org/10.1038/s41586-022-04411-y.

(2) Mullen, J. L.; Tsueng, G.; Latif, A. A.; Alkuzweny, M.; Cano, M.; Haag, E.; Zhou, J.; Zeller, M.; Hufbauer, E.; Matteson, N.; Andersen, K. G.; Wu, C.; Su, A. I.; Gangavarapu, K.; Hughes, L. D. https://outbreak.info/.

(3) Elbe, S.; Buckland-Merrett, G. Data, Disease and Diplomacy: GISAID’s Innovative Contribution to Global Health. Global Challenges 2017, 1 (1), 33–46. https://doi.org/10.1002/gch2.1018.

(4) Shu, Y.; McCauley, J. GISAID: Global Initiative on Sharing All Influenza Data – from Vision to Reality. Eurosurveillance 2017, 22 (13). https://doi.org/10.2807/1560-7917.ES.2017.22.13.30494.

(5) Peccia, J.; Zulli, A.; Brackney, D. E.; Grubaugh, N. D.; Kaplan, E. H.; Casanovas-Massana, A.; Ko, A. I.; Malik, A. A.; Wang, D.; Wang, M.; Warren, J. L.; Weinberger, D. M.; Omer, S. B. SARS-CoV-2 RNA Concentrations in Primary Municipal Sewage Sludge as a Leading Indicator of COVID-19 Outbreak Dynamics. Nature Biotechnology 2020, 38, 1164–1167.

(6) Feng, S.; Roguet, A.; McClary-Gutierrez, J. S.; Newton, R. J.; Kloczko, N.; Meiman, J. G.; McLellan, S. L. Evaluation of Sampling, Analysis, and Normalization Methods for SARS-CoV-2 Concentrations in Wastewater to Assess COVID-19 Burdens in Wisconsin Communities. ACS ES&T Water 2021, 1 (8), 1955–1965. https://doi.org/10.1021/acsestwater.1c00160.

(7) Kim, S.; Kennedy, L. C.; Wolfe, M. K.; Criddle, C. S.; Duong, D. H.; Topol, A.; White, B. J.; Kantor, R. S.; Nelson, K. L.; Steele, J. A.; Langlois, K.; Griffith, J. F.; Zimmer-Faust, A. G.; McLellan, S. L.; Schussman, M. K.; Ammerman, M.; Wigginton, K. R.; Bakker, K. M.; Boehm, A. B. SARS-CoV-2 RNA Is Enriched by Orders of Magnitude in Primary Settled Solids Relative to Liquid Wastewater at Publicly Owned Treatment Works. Environ. Sci.: Water Res. Technol. 2022. https://doi.org/10.1039/D1EW00826A.

(8) Fernandez-Cassi, X.; Scheidegger, A.; Bänziger, C.; Cariti, F.; Corzon, A. T.; Ganesanandamoorthy, P.; Lemaitre, J. C.; Ort, C.; Julian, T. R.; Kohn, T. Wastewater Monitoring Outperforms Case Numbers as a Tool to Track COVID-19 Incidence Dynamics When Test Positivity Rates Are High. Water Research 2021, 117252. https://doi.org/10.1016/j.watres.2021.117252.

(9) Lee, W. L.; Imakaev, M.; Armas, F.; McElroy, K. A.; Gu, X.; Duvallet, C.; Chandra, F.; Chen, H.; Leifels, M.; Mendola, S.; Floyd-O’Sullivan, R.; Powell, M. M.; Wilson, S. T.; Berge, K. L. J.; Lim, C. Y. J.; Wu, F.; Xiao, A.; Moniz, K.; Ghaeli, N.; Matus, M.; Thompson, J.; Alm, E. J. Quantitative SARS-CoV-2 Alpha Variant B.1.1.7 Tracking in Wastewater by Allele-Specific RT-QPCR. Environ. Sci. Technol. Lett. 2021, 8 (8), 675–682. https://doi.org/10.1021/acs.estlett.1c00375.

(10) Wolfe, M. K.; Hughes, B.; Duong, D.; Chan-Herur, V.; Wigginton, W. K.; White, B. J.; Boehm, A. B. Detection of SARS-CoV-2 Variants Mu, Beta, Gamma, Lambda, Delta, Alpha, and Omicron in Wastewater Settled Solids Using Mutation-Specific Assays Is Associated with Regional Detection of Variants in Clinical Samples. Applied and Environmental Microbiology 0 (0), e00045–22. https://doi.org/10.1128/aem.00045-22.

(11) Yaniv, K.; Ozer, E.; Kushmaro, A. SARS-CoV-2 Variants of Concern, Gamma (P.1) and Delta (B.1.617), Sensitive Detection and Quantification in Wastewater Employing Direct RT-QPCR. medRxiv 2021, 2021.07.14.21260495. https://doi.org/10.1101/2021.07.14.21260495.

(12) Crits-Christoph Alexander; Kantor Rose S.; Olm Matthew R.; Whitney Oscar N.; Al-Shayeb Basem; Lou Yue Clare; Flamholz Avi; Kennedy Lauren C.; Greenwald Hannah; Hinkle Adrian; Hetzel Jonathan; Spitzer Sara; Koble Jeffery; Tan Asako; Hyde Fred; Schroth Gary; Kuersten Scott; Banfield Jillian F.; Nelson Kara L.; Pettigrew Melinda M. Genome Sequencing of Sewage Detects Regionally Prevalent SARS-CoV-2 Variants. mBio 12 (1), e02703–20. https://doi.org/10.1128/mBio.02703-20.

(13) Yu, A.; Hughes, B.; Wolfe, M.; Leon, T.; Duong, D.; Rabe, A.; Kennedy, L.; Ravuri, S.; White, B.; Wigginton, K. R.; Boehm, A.; Vugia, D. Estimating Relative Abundance of Two SARS-CoV-2 Variants through Wastewater Surveillance at Two Large Metropolitan Sites. Emerging Infect. Dis. 2022, 28 (5), 940–947.

(14) Wolfe, M. K.; Topol, A.; Knudson, A.; Simpson, A.; White, B.; Duc, V.; Yu, A.; Li, L.; Balliet, M.; Stoddard, P.; Han, G.; Wigginton, K. R.; Boehm, A. High-Frequency, High-Throughput Quantification of SARS-CoV-2 RNA in Wastewater Settled Solids at Eight Publicly Owned Treatment Works in Northern California Shows Strong Association with COVID-19 Incidence. mSystems 2021, 0 (0), e00829–21. https://doi.org/10.1128/mSystems.00829-21.

(15) Topol, A.; Wolfe, M.; White, B.; Wigginton, K.; Boehm, A. High Throughput SARS-COV-2, PMMOV, and BCoV Quantification in Settled Solids Using Digital RT-PCR. protocols.io 2021.

(16) Kitajima, M.; Sassi, H. P.; Torrey, J. R. Pepper Mild Mottle Virus as a Water Quality Indicator. npj Clean Water 2018, 1 (1), 19. https://doi.org/10.1038/s41545-018-0019-5.

(17) Symonds, E. M.; Nguyen, K. H.; Harwood, V. J.; Breitbart, M. Pepper Mild Mottle Virus: A Plant Pathogen with a Greater Purpose in (Waste)Water Treatment Development and Public Health Management. Water Research 2018, 144, 1–12. https://doi.org/10.1016/j.watres.2018.06.066.

(18) McClary-Gutierrez, J. S.; Aanderud, Z. T.; Al-faliti, M.; Duvallet, C.; Gonzalez, R.; Guzman, J.; Holm, R. H.; Jahne, M. A.; Kantor, R. S.; Katsivelis, P.; Kuhn, K. G.; Langan, L. M.; Mansfeldt, C.; McLellan, S. L.; Mendoza Grijalva, L. M.; Murnane, K. S.; Naughton, C. C.; Packman, A. I.; Paraskevopoulos, S.; Radniecki, T. S.; Roman, F. A.; Shrestha, A.; Stadler, L. B.; Steele, J. A.; Swalla, B. M.; Vikesland, P.; Wartell, B.; Wilusz, C. J.; Wong, J. C. C.; Boehm, A. B.; Halden, R. U.; Bibby, K.; Delgado Vela, J. Standardizing Data Reporting in the Research Community to Enhance the Utility of Open Data for SARS-CoV-2 Wastewater Surveillance. Environ. Sci.: Water Res. Technol. 2021, 7 (9), 1545–1551. https://doi.org/10.1039/D1EW00235J.

(19) Borchardt, M. A.; Boehm, A. B.; Salit, M.; Spencer, S. K.; Wigginton, K. R.; Noble, R. T. The Environmental Microbiology Minimum Information (EMMI) Guidelines: QPCR and DPCR Quality and Reporting for Environmental Microbiology. Environ. Sci. Technol. 2021, 55 (15), 10210–10223. https://doi.org/10.1021/acs.est.1c01767.

(20) Topol, A.; Wolfe, M.; White, B.; Wigginton, K.; Boehm, A. High Throughput Pre-Analytical Processing of Wastewater Settled Solids for SARS-CoV-2 RNA Analyses. protocols.io 2021.

