## Supplemental Information for "Regional replacement of SARS-CoV-2 variant BA.1 with BA.2 as observed through wastewater surveillance"

### Supporting information
#### For

**Regional replacement of SARS-CoV-2 variant BA.1 with BA.2 as observed through wastewater surveillance**

Alexandria B. Boehm[1]*, Bridgette Hughes[2], Marlene K. Wolfe[3], Bradley J. White[2], Dorothea Duong[2], Vikram Chan-Herur[2]

1. Department of Civil & Environmental Engineering, Stanford University, 473 Via Ortega, Stanford, CA, USA, 94305
2. Verily Life Sciences, South San Francisco, CA, USA, 94080
3. Gangarosa Department of Environmental Health, Rollins School of Public Health, Emory University, 1518 Clifton Rd, Atlanta, GA, USA, 30322

**Number of pages: 12**
**Number of figures: 3**
**Number of tables: 5**

#### Materials and Methods

**Assay specificity and sensitivity testing.** Primers and probe sequences (Table 1) were screened for specificity in silico using NCBI Blast, and then tested in vitro against a virus panel (NATtrol™ Respiratory Verification Panel, NATRVP2-BIO, Zeptomatrix) that includes several influenza and coronavirus viruses, "wild-type" gRNA from SARS-CoV-2 strain 2019-nCoV/USA-WA1/2020 (ATCC® VR-1986D™) which does not contain the mutations (hereafter referred to as WT-gRNA) as well as a combination of heat inactivated SARS-CoV-2 stain B.1.1.7 (SARS-CoV-2 variant B.1.1.7, ATCC® VR-3326HK™), a positive clinical sample confirmed as Mu provided by Dr. Ben Pinsky at Stanford Virology Laboratory, and synthetic gRNA from Twist Biosciences (South San Francisco, California, USA) for Beta (Twist control 16), Gamma (Twist control 17), Delta (Twist control 23), Omicron BA.1 (Twist control 48), and Omicron BA.2 (Twist control 50) (Table S1). RNA was extracted from the virus panel and whole viruses using the Perkin Elmer Chemagic Viral RNA extraction kit (Chemagic Kit CMG-1033-S designed for SARS-CoV-2). RNA was used undiluted as template in digital droplet PCR with mutation primer and probes. The concentration of gRNA used in the in vitro specificity testing was approximately 275 copies per well. The mutation assays were challenged against the respiratory panel gRNA in single wells, and non-target variant gRNA in 8 replicate wells. PCR plates contained positive controls and negative PCR no template controls (NTC) in 8 wells (Table S1).

The sensitivity and specificity of the mutation assays were further tested by diluting target variant gRNA (Table S1) in no (0 copies), low (100 copies), and high (10,000 copies) background of WT SARS-CoV-2 gRNA. For the assay targeting LPPA24S, the Twist synthetic gRNA of BA.2 (Twist control 50) was used as target gRNA. For the S:477-505 mutation assay, an equal molal mixture of Twist synthetic gRNA of BA.1 (Twist control 48) and BA.2 (Twist control 50) was used as target gRNA. Each dilution was run in three replicate wells. The number of copies of variant mutation sequences input to each well was estimated using a dilution series of variant gRNA in no background; the vendor specified concentration of the variant gRNA was scaled by the slope of the curve relating the measured ddRT-PCR concentration and the calculated input concentration based on the vendor estimates. Our experience suggests vendor estimates can be imprecise. PCR negative controls were run in 4 wells per plate.

**Wastewater sampling collection.** Samples were collected by POTW staff using sterile technique in clean, labeled bottles provided by our team as part of a long term wastewater surveillance project. Approximately 50 mL of settled solids was collected each day from each sewage treatment plant between 1 January 2022 and 12 or 13 April 2022. At seven of the eight POTWs, settled solids were collected from the primary clarifier; at these POTWs, the residence time of solids in the primary clarifier ranged from 1 to 6 h. Settled solids samples were grab samples at all plants except for SJ. At SJ, POTW staff manually collected a 24-h composite sample. At Gil, solids were settled from a 24-h composite influent sample using standard method 160.5 [1]. Samples were immediately stored at 4°C and transported to the commercial partner laboratory by a courier service where processing began within 6 h of collection.

**RNA extraction and ddRT-PCR.** RNA was extracted from the 10 replicate aliquots of dewatered settled solids using the Chemagic Viral DNA/RNA 300 kit H96 for the Perkin Elmer Chemagic 360 (Perkin Elmer, Waltham, MA) followed by PCR inhibitor removal with the Zymo OneStep-96 PCR Inhibitor Removal kit (Zymo Research, Irvine, CA) as described in Wolfe et al.[2–4] ddRT-PCR was performed on 20 µl samples from a 22 µl reaction volume, prepared using 5.5 µl template, mixed with 5.5 µl of One-Step RT-ddPCR Advanced Kit for Probes (Bio-Rad 1863021), 2.2 µl of 200 U/µl Reverse Transcriptase, 1.1 µl of 300 mM DTT and primers and probes at a final concentration of 900 nM and 250 nM respectively. Primer and probes were purchased from Integrated DNA Technologies (IDT, San Diego, CA).

Droplets were generated using the AutoDG Automated Droplet Generator (Bio-Rad, Hercules, CA). PCR was performed using Mastercycler Pro (Eppendforf, Enfield, CT) with with the following cycling conditions: reverse transcription at 50°C for 60 minutes, enzyme activation at 95°C for 5 minutes, 40 cycles of denaturation at 95°C for 30 seconds and annealing and extension at 61°C for 30 seconds, enzyme deactivation at 98°C for 10 minutes then an indefinite hold at 4°C. The ramp rate for temperature changes were set to 2°C/second and the final hold at 4°C was performed for a minimum of 30 minutes to allow the droplets to stabilize. Droplets were analyzed using the QX200 Droplet Reader (Bio-Rad). A well had to have over 10,000 droplets for inclusion in the analysis. All liquid transfers were performed using the Agilent Bravo (Agilent Technologies, Santa Clara, CA).

For wastewater samples, positive PCR controls for each target assayed on the plate were run in 1 well, PCR NTCs in three wells, extraction negative controls (where no sample was added to the extraction kit) in three wells, and one extraction positive control (containing SARS-CoV-2 gRNA) in one well. PCR positive controls consisted of gRNA of SARS-CoV-2 (strain 2019-nCoV/USA-WA1/2020, ATCC® VR-1986D™) as well as BA.1 and BA.2 (Twist synthetic control gRNA 48 and 50 ).

Table S1. Details of variant assays used in this study. Details on fraction of variants containing mutation obtained from outbreak.info on 16 April 2022. * after the Pango lineage BA.1 and BA.2 indicates all sub lineages (19 and 12, respectively) of the associated lineage.

| Omicron Variant (s) | Mutation(s) located on S gene | Fraction of variant containing mutation (# containing mutation / total #) | Fraction of non-target variant containing mutation | Positive control RNA using in sensitivity testing | RNA used to test specificity (negative controls) |
|---|---|---|---|---|---|
| Omicron BA.1* | del143-145 | 93% (2032186/ 2177814) | BA.2= 0% (0 / 2177814) | Described in Wolfe et al.[5] | Described in Wolfe et al.[5] |
| Omicron BA.2* | LPPA24S (del25-27,L24S) | 92% (759428/ 822506) | BA.1 = 0.003% (28 / 822506) | Synthetic gRNA from TWIST control 50 (BA.2) | WT, gRNA, Alpha, Beta, Gamma, Delta, Mu, Lambda, BA.1 |
| Omicron (BA.1*, BA.2*) | S477N, T478K, Q493R , Q498R, Y505H | 85% (2554632/ 3000302) | NA | Synthetic gRNA from TWIST controls 48 & 50 (BA.1 and BA.2) | WT, gRNA, Alpha, Beta, Gamma, Delta, Mu, Lambda, |

Table S2. The parameters used in assay development (that controlled sequence length, GC content, and melt temperatures) in Primer3Plus (https://primer3plus.com/).

- Product size ranges: 60-200
- Primer size: min 15, opt 20, max 36
- Primer melting temperature: min 50°C, optimal 60°C, max 65°C
- GC% content: min 40%, optimal 50%, high 60%
- concentration of divalent cations = 3.8 mM
- concentration of dNTPs needs to be 0.8 mM
- Internal Oligo: size min 15, optimal 20, max 27
- Internal Oligo: Melting temp min 62°C, optimal 63°C, max 70°C
- Internal Oligo: GC% min 30%, optimal 50%, max 80%

Table S3. The date in 2022 on which LPPA24S/N became larger than del143-145/N; date identified using 5-d trimmed average lines in Figure 2; using a sigmoid function to fit the relative mutation time series and then using those fits to identify the date did not change the date.

| POTW | Date |
|---|---|
| PA | 4 March |
| SJ | 10 March |
| Gil | 19 March |
| Sun | 6 March |
| SVCW | 10 March |
| Ocean | 26 Feb |
| Dav | 14 March |
| Sac | 19 March |

Table S4. Kendall’s tau, p-values, and number of paired measurements. Top panel: Values represent results from Kendall’s tau test of association between relative concentrations of del143-145 (both 5-d trimmed average and raw values) and the proportion of clinical specimens in California assigned to BA.1*. Bottom panel: Values represent results from Kendalls tau test of association between relative concentrations of LPPA24S (both 5-d trimmed average and raw values) and the proportion of clinical specimens in California assigned to BA.2*.

| | 5-d trimmed average relative concentration del143-145 | | | raw relative concentration del143-145 | | |
|---|---|---|---|---|---|---|
| POTW | tau BA.1* | p value BA.1* | n BA.1* | tau BA.1* | p value BA.1* | n BA.1* |
| PA | 0.68 | 8.1E-15 | 101 | 0.56 | 9.8E-10 | 101 |
| SJ | 0.59 | 9.8E-11 | 102 | 0.48 | 3.0E-07 | 102 |
| Sun | 0.61 | 5.7E-12 | 103 | 0.56 | 5.7E-10 | 103 |
| Gil | 0.51 | 5.2E-08 | 102 | 0.38 | 8.7E-05 | 102 |
| SVCW | 0.69 | 1.2E-15 | 103 | 0.55 | 1.4E-09 | 103 |
| Ocean | 0.73 | 7.2E-17 | 94 | 0.65 | 1.8E-12 | 94 |
| Sac | 0.81 | 2.4E-25 | 103 | 0.65 | 1.1E-13 | 103 |
| Dav | 0.70 | 1.8E-16 | 103 | 0.54 | 4.8E-09 | 101 |

| | 5-d trimmed average relative concentration LPPA24S | | | raw average relative concentration LPPA24S | | |
|---|---|---|---|---|---|---|
| POTW | tau BA.2* | p value BA.2* | n BA.2* | tau BA.2* | p value BA.2* | n BA.2* |
| PA | 0.70 | 4.5E-10 | 61 | 0.63 | 5.6E-08 | 61 |
| SJ | 0.89 | 1.3E-21 | 62 | 0.78 | 1.0E-13 | 62 |
| Sun | 0.87 | 4.0E-20 | 62 | 0.69 | 7.9E-10 | 62 |
| Gil | 0.67 | 4.4E-09 | 61 | 0.54 | 8.6E-06 | 61 |
| SVCW | 0.76 | 4.8E-13 | 62 | 0.62 | 6.3E-08 | 62 |
| Ocean | 0.73 | 8.2E-11 | 58 | 0.64 | 1.1E-07 | 57 |
| Sac | 0.90 | 8.5E-24 | 62 | 0.72 | 5.3E-11 | 62 |
| Dav | 0.74 | 3.9E-12 | 62 | 0.60 | 4.7E-07 | 60 |

Table S5. Primers and probes used in this study that have been previously published by Wolfe et al.[2] Information on the fluorescent molecule and quenchers used for the probes are provided in parentheses after their sequence. FAM, 6-fluorescein amidite; HEX, hexachloro-fluorescein; ZEN, a proprietary internal quencher from IDT; IBFQ, Iowa Black FQ.

| | | |
|---|---|---|
| N Gene | Forward | CATTACGTTTGGTGGACCCT |
| | Reverse | CCTTGCCATGTTGAGTGAGA |
| | Probe | CGCGATCAAAACAACGTCGG (5' FAM/ZEN/3' IBFQ) |
| Del143-145 | Forward | ATTCGAAGACCCAGTCCCTA |
| | Reverse | ACTCTGAACTCACTTTCCATCC |
| | Probe | TTGTAATGATCCATTTTTGGACCACAA(5' FAM or HEX/ZEN/3' IBFQ) |
| BCoV | Forward | CTGGAAGTTGGTGGAGTT |
| | Reverse | ATTATCGGCCTAACATACATC |
| | Probe | CCTTCATATCTATACACATCAAGTTGTT (5' FAM/ZEN/3' IBFQ) |
| PMMoV | Forward | GAGTGGTTTGACCTTAACGTTTGA |
| | Reverse | TTGTCGGTTGCAATGCAAGT |
| | Probe | CCTACCGAAGCAAATG (5' HEX/ZEN/3' IBFQ) |

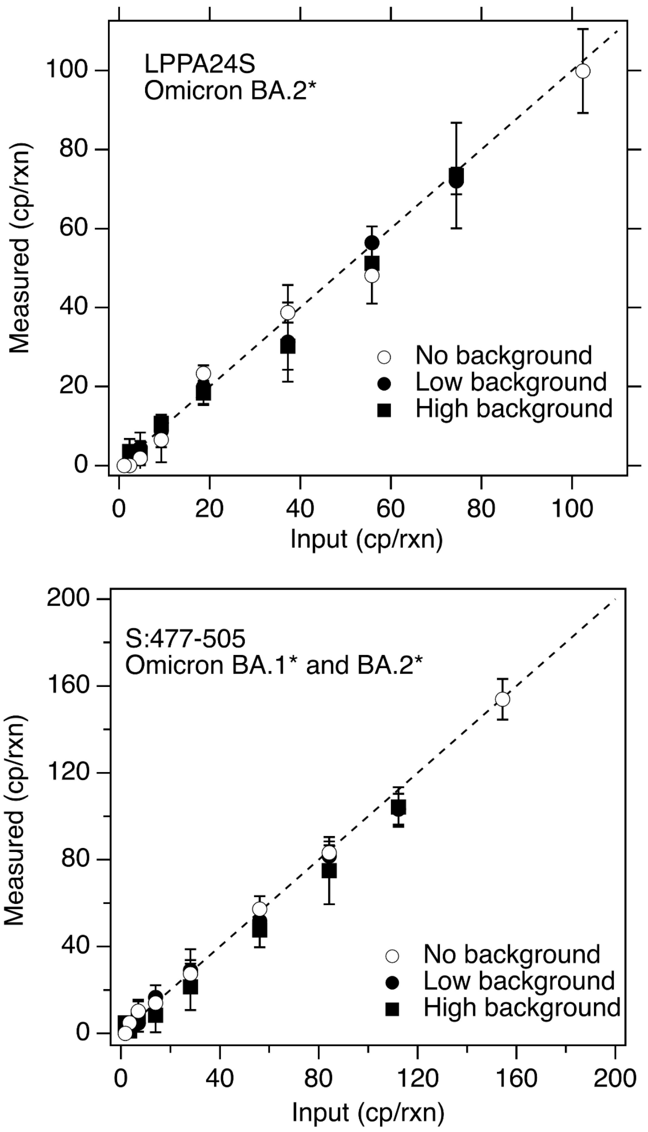


Figure S1. Copies (cp) of mutations measured per reaction (rxn) when RNA containing the mutation was diluted into no, low, and high background of WT-gRNA. Low background is 100 copies/well and high background is 10,000 copies/well where “copies” refers to copies of genomes of WT-gRNA. Markers show average across three replicate wells and error bars represent standard deviations. In some cases, the error bar is not visible because it is smaller than the marker.

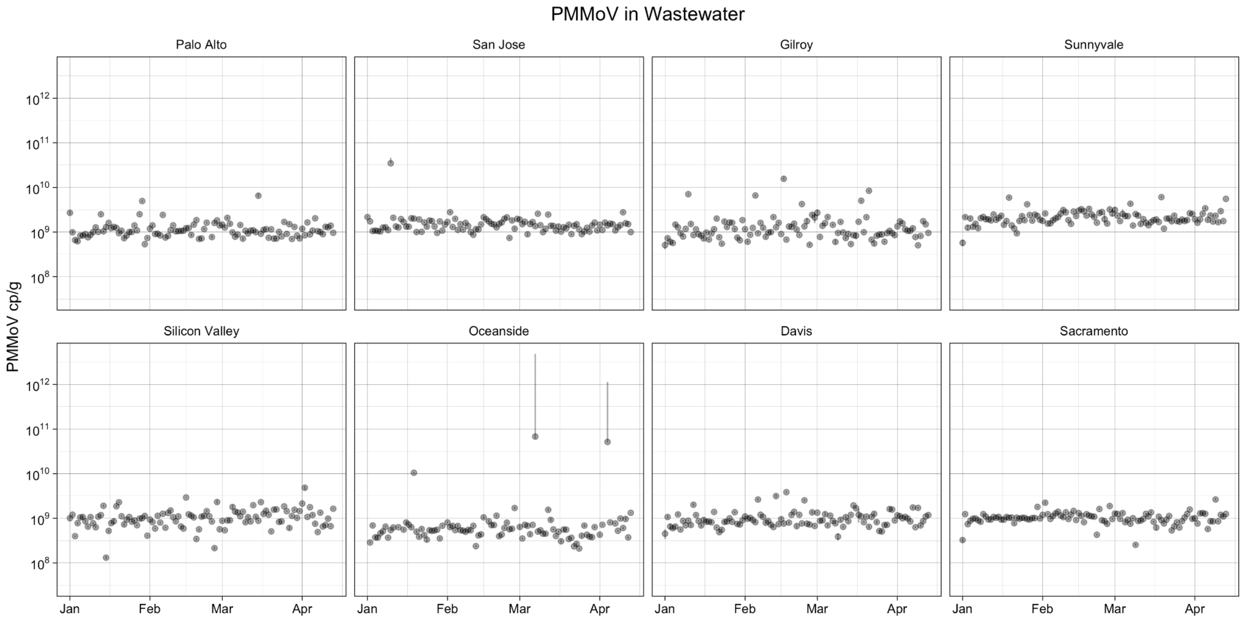


Figure S2. Pepper Mild Mottle Virus (PMMoV) concentrations at 8 POTWs in the study (name indicated in corner). Error bars are 68% confidence intervals that include error from variation among 10 replicate wells, and Poisson error ("total error" as reported by the instrument). If error bars cannot be seen, they are smaller than the symbol.

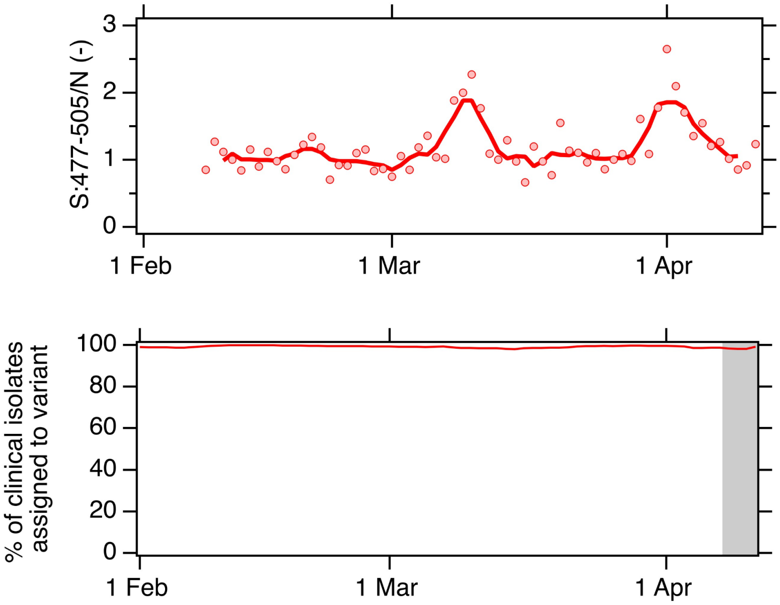


Figure S3. Top panel: Relative concentration of S:477-505; raw data are shown as markers, and line represents 5-d trimmed average. Bottom panel: Percentage of clinical cases in California classified as BA.1* and BA.2* as a function of specimen collection date; data represent 7-d moving average as acquired from GISAID. Data available through 13 April when downloaded on 21 April 2022. Grey area represents time period for which clinical sequence data were not yet available when the last wastewater measurement was made public on our website (wbe.stanford.edu).

References


(1) AWWA. *Standard Methods for the Examination of Water and Wastewater*, 21st ed.; Eaton, A. D., Clesceri, L. S., Rice, E. W., Greenberg, A. E., Series Eds.; American Public Health Association, American Water Works Association, Water Environment Federation: Baltimore, 2005.

(2) Wolfe, M. K.; Topol, A.; Knudson, A.; Simpson, A.; White, B.; Duc, V.; Yu, A.; Li, L.; Balliet, M.; Stoddard, P.; Han, G.; Wigginton, K. R.; Boehm, A. High-Frequency, High-Throughput Quantification of SARS-CoV-2 RNA in Wastewater Settled Solids at Eight Publicly Owned Treatment Works in Northern California Shows Strong Association with COVID-19 Incidence. *mSystems* **2021**, *0* (0), e00829-21. https://doi.org/10.1128/mSystems.00829-21.

(3) Topol, A.; Wolfe, M.; Wigginton, K.; White, B.; Boehm, A. High Throughput RNA Extraction and PCR Inhibitor Removal of Settled Solids for Wastewater Surveillance of SARS-CoV-2 RNA. *protocols.io* **2021**.

(4) Topol, A.; Wolfe, M.; White, B.; Wigginton, K.; Boehm, A. High Throughput Pre-Analytical Processing of Wastewater Settled Solids for SARS-CoV-2 RNA Analyses. *protocols.io* **2021**.

(5) Wolfe, M. K.; Hughes, B.; Duong, D.; Chan-Herur, V.; Wigginton, W. K.; White, B. J.; Boehm, A. B. Detection of SARS-CoV-2 Variants Mu, Beta, Gamma, Lambda, Delta, Alpha, and Omicron in Wastewater Settled Solids Using Mutation-Specific Assays Is Associated with Regional Detection of Variants in Clinical Samples. *Applied and Environmental Microbiology 0* (0), e00045-22. https://doi.org/10.1128/aem.00045-22.
